# Personalising Transcranial Magnetic Stimulation Therapy for Neuropathic Pain with Somato-Cognitive Action Network Connectivity to Cingulo-Opercular Network: A Preliminary Open-Label Study

**DOI:** 10.64898/2026.08.23.26361115

**Authors:** Zhimin Huang, Han Li, Yunze Li, Shengqiao Wang, Andrew Zalesky, Robin Cash, Xianwei Che, Zhiying Feng

## Abstract

**Background:** Neuropathic pain (NP) remains a therapeutic challenge, with conventional repetitive transcranial magnetic stimulation (rTMS) of the primary motor cortex (M1) yielding a response rate of approximately 40%. Personalised targeting based on dysfunctional neurocircuitry offers a promising strategy to enhance efficacy, yet its application in NP is unexplored. This open-label trial investigated a novel targeting approach guided by the recently described cingulo-opercular and somato-cognitive action (CON-SCAN) network, a circuit integrating cognitive and affective dimensions of pain.

**Methods:** Twenty patients with NP received 10 sessions of M1-rTMS over two weeks, with the stimulation site individually localised based on maximal functional connectivity to a CON template.

**Results:** Increased CON-SCAN connectivity from baseline to post-treatment was associated with reduction in pain interference, anxiety and depression scores. The response rate was 50% post-treatment, which was maintained at the 1-month follow-up. Improvements were also observed in neuropathic pain symptoms, negative affect, and overall health.

**Conclusions:** As the first connectivity-guided rTMS trial for NP, this study provides preliminary evidence that personalised targeting of the CON-SCAN network is feasible and associated with the analgesic effects of M1-rTMS, supporting further investigation in randomised controlled trials.

**Trial registration:** Chinese Clinical Trial Registry, ChiCTR2400094568. Registered 24 December 2024, http://www.chictr.org.cn.

## BACKGROUND

Neuropathic pain (NP), a refractory condition affecting nearly 10% of the global population, remains a therapeutic challenge with first-line medications often limited by side effects (1–3). Non-invasive brain stimulation (NIBS) has emerged as a promising therapeutic approach, with repetitive transcranial magnetic stimulation (rTMS) over the primary motor cortex (M1) constituting a safe and effective treatment (4–6). However, the clinical application of rTMS in neuropathic pain is limited by treatment response, generally defined as ≥ 30% in pain reduction (7, 8). A commonly used rTMS protocol for neuropathic pain is 10 sessions over 2 weeks (10-Hz, ∼1,500 pulses per session), which tends to yield ∼40% response rate (9–11).

A key strategy for enhancing rTMS efficacy is to optimise stimulation targets to engage disorder-specific circuits. Exemplifying this in depression, superior clinical outcomes are achieved when rTMS is applied to regions of the dorsolateral prefrontal cortex (DLPFC) that are functionally connected to a depression circuit centred on the subgenual cingulate cortex (SGC), a key node in mood regulation (12–15). In the context of neuropathic pain, two independent reviews have urged the development and implementation of personalised targeting strategies to enhance treatment outcomes (16, 17). Unfortunately, there has been no study to date to investigate the efficacy of personalised rTMS treatment for neuropathic pain.

For decades, the hand knob of the primary motor cortex (M1) has served as the most common and empirically validated conventional target for TMS pain therapy. The robust analgesic effects of M1-rTMS are widely considered to rely on broadly distributed network mechanisms rather than being strictly reducible to localized motor representations. Building upon on this evidence, recent advances in functional connectomics offer an opportunity to further optimize this approach by identifying the precise intra-cortical circuits mediating these distributed effects (18).

Critically, it was recently demonstrated that the classic motor homunculus is punctuated by “inter-effector” regions, which are not involved in motor action. These regions have with strong connectivity to each other, and to the cingulo-opercular network (CON), which is critical for a range of functions including pain (18). The hand region, traditionally targeted by TMS, is positioned between two such inter-effector regions, raising the intriguing possibility that the established efficacy of conventional hand knob targeting may, in part, be driven by the recruitment of these adjacent inter-effector regions. Consequently, intentionally and prospectively targeting these regions may provide a circuit-informed refinement to enhance therapeutic outcomes. Collectively the inter-effector system has been termed the somato-cognitive action network (SCAN). The CON network, with which the SCAN is closely associated, contains regions that are associated with pain including anterior cingulate cortex (ACC), insula, and frontal regions (19). TMS response in pain has been associated with modulation of the activity and connectivity of these regions(20), which are located downstream of the stimulation site (4, 21). These regions are widely accepted to encode the attentional cognitive (e.g. frontal regions, insula) or emotional affective (e.g. ACC) dimensions of pain (22).

We conducted a prospective clinical study to investigate the therapeutic efficacy of CON-SCAN network guided the personalised rTMS treatment for neuropathic pain. TMS targets were constrained to a spatial mask delineating the motor cortical bounds of SCAN, and optimised according to person-specific connectivity with CON, using a cluster-based targeting approach adapted from the depression literature (23). A course of 10 treatment sessions was then delivered over 2 weeks to neuropathic participants (n = 20). It was hypothesised that personalised rTMS treatment would increase treatment response for neuropathic pain compared to a 40% response rate in the literature. The primary outcome measure was pain intensity assessed using the Visual Analogue Scale, with a treatment response defined as a ≥30% reduction in pain scores from baseline (8, 24).

## METHODS

### 2.1 Study overview

This prospective, open-label trial was registered in the Chinese Clinical Trial Registry (ChiCTR2400094568, registered on December 24, 2024; http://www.chictr.org.cn). This study received ethical approval from the Ethics Committees of the Affiliated Hospital of Hangzhou Normal University (2024-E2-HS-008) and was conducted in accordance with the Declaration of Helsinki. All participants provided written informed consent prior to participation. Within 2 weeks, patients received 10 sessions (interval ≥ 24 hours) of 10-Hz repetitive transcranial magnetic stimulation (rTMS). Clinical assessments were conducted at three time points: before the intervention (Baseline), immediately after the final treatment session (Post), and one month following the final session (Follow-up). In addition, structural and functional MRI scans were acquired at both Baseline and Post. All rTMS procedures were administered by an operator (Z.H.) who was not involved in clinical assessments. Meanwhile, all clinical assessments were performed by a trained neurologist (H.L.).

### 2.2 Participants

The sample size was calculated using G*Power software (version 3.1) (25). The calculation was based on an a priori power analysis for a two-tailed paired t-test to detect within-subject changes from baseline to post-treatment. Based on our preliminary data (Baseline: 4.98±1.97; Post-treatment: 2.28±2.51), the correlation between baseline and post-treatment measures was assumed to be 0.3, yielding a calculated Cohen’ s d_z_ of 1.00(26). To minimize the risk of Type II errors, a stringent statistical power (1−β) of 0.90 was adopted. With the significance level being set at 0.05, power analysis indicated that a minimum of 13 participants would be required to detect the anticipated effect size. To ensure the robustness of our results and account for an anticipated dropout rate of 20%, we ultimately recruited 20 participants, ensuring the final sample remained sufficient to maintain adequate power.

Patient recruitment was conducted at the First Affiliated Hospital, Zhejiang University School of Medicine. A total of thirty-two individuals with chronic neuropathic pain were screened for eligibility in this study. The inclusion criteria were as follows: (1) a clinical diagnosis of neuropathic pain by the International Association for the Study of Pain (IASP) criteria, with a symptom duration of ≥3 months, or neuropathic pain components secondary to nociceptive pain (neuropathic pain screening tool: ID Pain score ≥2); (2) age between 18 and 75 years; (3) an average daily pain intensity of >30 mm on the Visual Analog Scale (VAS) during the week preceding enrolment; (4) a stable analgesic regimen with no dosage adjustments throughout the 1-week baseline assessment period; (5) physical and cognitive ability to undergo all study procedures, including clinical evaluations, MRI, and rTMS; and (6) capacity to provide written informed consent, demonstrating understanding of the study’s objectives, and willingness to adhere to the protocol.

The exclusion criteria were: (1) any contraindications to TMS or MRI (e.g., metallic implants, claustrophobia, inability to maintain a supine position); (2) psychiatric disorders or significant cognitive impairment, defined as a score >24 on the 17-item Hamilton Depression Rating Scale (HAMD-17), >29 on the 14-item Hamilton Anxiety Rating Scale (HAMA-14), or ≤24 on the Mini-Mental State Examination (MMSE); (3) severe cardiopulmonary dysfunction or general debilitation that would compromise cooperation with the study protocol; (4) other significant concurrent medical conditions that could interfere with study participation or interpretation of results (e.g., active epilepsy, recent myocardial infarction); (5) pregnancy or lactation; or (6) current participation in any other interventional clinical trial.

### 2.3 Computation of personalized treatment target

#### 2.3.1 MRI data acquisition

MRI data were acquired using a 3.0 Tesla GE Architect scanner at the Affiliated Hospital of Hangzhou Normal University. High-resolution T1-weighted structural images were obtained with a 3D magnetization-prepared rapid gradient echo (MP-RAGE) sequence using the following parameters: FOV = 256 mm, slice thickness = 1.0 mm, matrix = 256 × 256, TI = 1100 ms, flip angle = 7°, and receiver bandwidth = 31.25 kHz. A total of 192 slices were acquired in a single slab without gap.

Resting-state functional images were collected using a 2D gradient-echo echo-planar imaging (EPI) sequence with the following parameters: TR = 1000 ms, TE = 30 ms, flip angle = 60°, FOV = 216 mm, slice thickness = 3.0 mm, matrix = 72 × 72, 50 axial slices acquired in interleaved order, no slice gap. Acquisition was accelerated using a multiband factor of 3 combined with ARC parallel imaging. A total of 600 volumes were acquired over a 10-minute scan duration.

For EPI distortion correction, B_0_ field maps were acquired using a dual-echo gradient-echo sequence with the following parameters: TR = 400 ms, TE_1_ = 4.9 ms, TE_2_ = 7.4 ms, flip angle = 60°, FOV = 216 mm, matrix = 64 × 64, slice thickness = 3.0 mm, 50 slices acquired contiguously, and a receiver bandwidth of 31.25 kHz.

#### 2.3.2 Imaging preprocessing

All functional and structural MRI data were preprocessed using FSL version 6.0.0 (FMRIB Software Library, Oxford, UK) in combination with custom MATLAB R2022b (MathWorks, Natick, MA) scripts. Prior to processing in FSL, individual T1-weighted structural images were manually reoriented in SPM12 to set the image origin at the anterior commissure (AC) to facilitate subsequent co-registration. Functional MRI preprocessing comprised multiple stages: removal of the first 10 volumes for signal equilibrium; spatial and gradient distortion correction; head motion realignment using rigid-body transformation; co-registration of functional images to the corresponding T1-weighted structural images using boundary-based registration (BBR); intensity normalization across timepoints; and nonlinear spatial normalization to the MNI152 2mm template using FSL’s standard registration tools (FNIRT). Temporal high-pass filtering (cutoff: 0.01 Hz) was applied, followed by automated denoising using the FIX+ICA pipeline. To rigorously minimize physiological artifacts, mean signals from the white matter and cerebrospinal fluid were regressed out as nuisance covariates. However, Global Signal Regression (GSR) was not performed in order to avoid the artificial induction of negative correlations. Furthermore, no frame censoring (scrubbing) was applied, meaning that all 590 post-equilibrium volumes were retained for target computation. Head motion was strictly monitored. Recognizing the inherent challenges of scanning patients with chronic pain, the exclusion criterion for excessive motion was set at a mean relative displacement (frame-to-frame) greater than 0.4 mm. All 20 participants met this standard (cohort mean relative displacement = 0.10 ± 0.07 mm; maximum observed mean relative displacement across all scans = 0.38 mm), resulting in no exclusions. Finally, spatial smoothing was applied with a 4 mm full width at half-maximum (FWHM) Gaussian kernel.

#### 2.3.3 Region of interest

The cingulo-opercular network (CON) mask served as the seedmap for functional connectivity analysis. The optimal person-specific stimulation target was computed within the spatial bounds of the somato-cognitive action network (SCAN) mask using an established cluster-based approach (27). The target and seed networks were defined using pre-existing masks derived from large dataset (Human Connectome Project, HCP) (28).

#### 2.3.4 Computation of CON time series

The CON time series was defined using the seedmap approach rather than a conventional binary mask (28). This method was specifically chosen to maximise the signal-to-noise ratio (SNR) by leveraging data from all grey matter voxels instead of a limited number of voxels within a circumscribed ROI. As established by Fox et al, computing the time series as a weighted spatial average across the whole brain effectively suppresses stochastic noise and local artifacts, providing a more reliable and robust estimation of the network intrinsic activity (29). Specifically, the contribution of each voxel was weighted according to its group-averaged functional connectivity with the CON, derived from 1200 scans (two scans per individual, 600 individuals) in the HCP dataset (https://db.humanconnectome.org/).

#### 2.3.5 Computation of personalized SCAN treatment target

We used the cluster-based approach to identify the optimal target coordinates within the SCAN network (27). Specifically, individualised SCAN targets were determined via an iterative thresholding and clustering algorithm. We systematically varied the retention rate of top-connected voxels (from 0.1% to 50%) until a cluster of at least 26 contiguous voxels was identified. In a 3D grid, 26 voxels essentially constitute a complete spatial neighborhood (a 3×3×3 architecture) around a central peak. This stringent size filters out single-voxel noise while maintaining a highly focal ’epicenter’, allowing for maximum anatomical precision when computing the center-of-gravity target. By adopting the most stringent threshold that met this volume criterion, we ensured that the target was localised to the participant’s most significant functional peak. The centre of gravity of this cluster served as the final coordinate for neuronavigated rTMS(Figure 1). Crucially, while the network templates and initial peak connectivity searches were computed in standard MNI152 space, the final optimal coordinate for each participant was transformed back into their individual native T1 structural space. This reverse transformation was executed using the inverse non-linear warp fields derived from the FNIRT normalization process. The resulting native-space coordinates were then directly imported into the Brainsight neuronavigation system(v2.5.1, Rogue Research Inc., Montreal, QC, Canada) alongside the native T1 images. To ensure alignment accuracy, we visually verified that each functional target was correctly localized on the precentral gyrus using the 3D-reconstructed brain surface within the Brainsight interface.This procedure ensured perfect spatial alignment between the EPI-derived functional targets and the real-time structural guidance utilized during the rTMS sessions.

**Figure 1.**
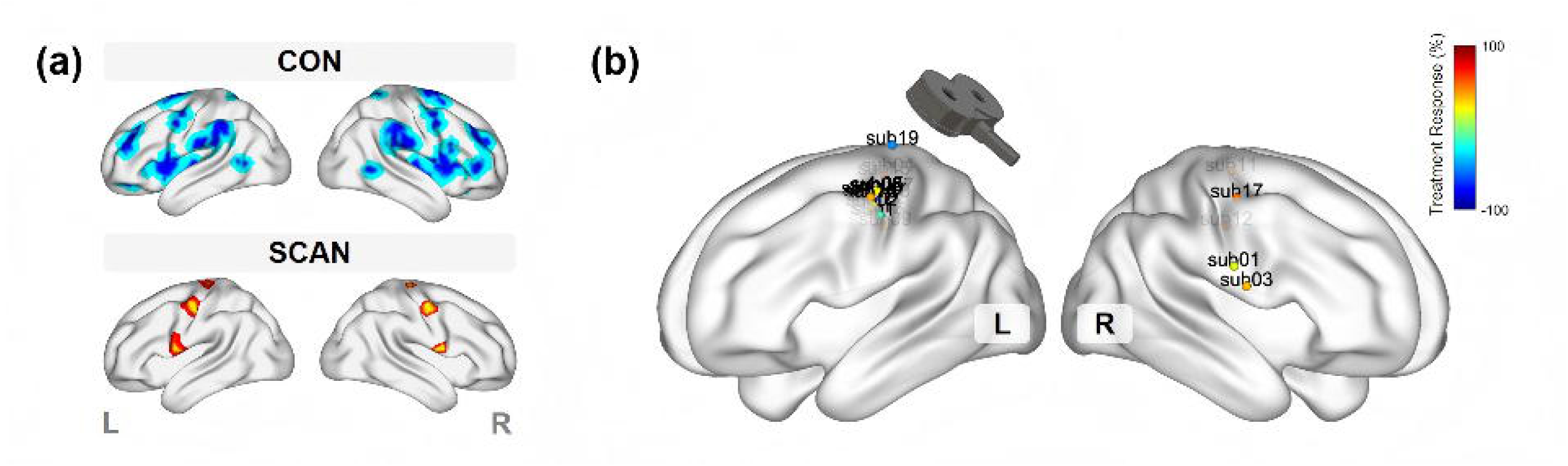
Target Network Mapping and Individualized Coordinate Presentation. (a)Anatomical Location of Target Networks. Spatial overlay of the Somato-Cognitive Action Network (SCAN; red) and Cingulo-Opercular Network (CON; blue) masks, defining the regions of interest (ROIs) for functional connectivity analyses. (b) Individualized Stereotactic Coordinates.

Finally, to evaluate the spatial dispersion of the individualized stimulation targets across the cohort, all target coordinates were mathematically mirrored to the left hemisphere (by taking the negative absolute value of the X-coordinate) to account for stimulation laterality. We then computed the group-average centroid of these mirrored coordinates and calculated the Euclidean distance between each participant’s individualized target and the group centroid.

### 2.4 Change in SCAN-CON functional connectivity following treatment

To understand whether changes in pain following TMS therapy relate to changes in SCAN-CON functional connectivity, we computed the representative time series of each network. The CON time series was extracted following the procedure detailed in Section 2.3.4. For the SCAN, we averaged the BOLD time courses across all constituent voxels within a binary network mask to generate a single representative signal. While analyzing the specific stimulation target provides a localized view of treatment effects, we employed this network-averaging approach to capture the coherent oscillations and functional integration of the SCAN as a whole. Given that chronic pain is a complex, systemic state, we hypothesized that clinical improvement would be more closely linked to the overall coherence and communication between large-scale networks (SCAN-CON) rather than isolated changes at the local stimulation site. Furthermore, this network-level signal-to-noise ratio (SNR) is inherently higher than that of individual regions-of-interest, providing greater sensitivity for detecting associations with behavioral and clinical outcomes.

Functional connectivity between the SCAN and CON network was quantified as the Pearson correlation coefficient between their respective mean time series. This computation was implemented in MATLAB (MathWorks, Natick, MA, USA) using the built-in corr function, yielding a scalar correlation value per participant that served as the connectivity metric.

### 2.5 rTMS treatment protocol

The enrolled patients completed 10 rTMS sessions over 2 weeks. All treatments were administered using a Magstim Rapid^2^ stimulator (Magstim Company Ltd, UK) with a figure-eight coil, guided by a neuronavigation system (Brainsight 2.3; Rogue Research, Canada) for precise and consistent targeting. Individual T1-weighted structural images were acquired and reconstructed into 3D brain models to facilitate targeting. The centre of the coil was placed over the personalised target in the M1 area (specific coordinates are detailed in Section 2.3.5), targeting the hemisphere contralateral to the painful side. Specifically, the laterality of the stimulation target was determined according to the following predefined criteria: for strictly unilateral pain, the contralateral hemisphere was targeted. In cases of bilateral, multifocal, or poorly lateralized pain, stimulation was delivered contralateral to the predominantly painful side. For patients experiencing symmetrical bilateral or centralized midline pain, the left hemisphere was uniformly selected as the default therapeutic target. (See supplementary Table 1)

The stimulation parameters were set as follows: a frequency of 10 Hz, delivered in 5-second trains separated by 25-second intervals, resulting in a total of 36 trains and 1800 pulses per session (30, 31). The stimulation intensity was set at 90% of the individual’s resting motor threshold (RMT) (11, 32, 33). The RMT was defined as the minimum intensity required to elicit motor-evoked potentials (MEPs) greater than 0.05 mV in at least 5 out of 10 trials. MEPs were continuously monitored and recorded from the first dorsal interosseous muscle using surface electrodes (Kendall ECG Long-Term Monitoring Electrodes, H124SG) and an integrated electromyography device.

### 2.6 Outcome measures

The outcome measures were reported according to the IMMPACT recommendations for clinical trials in chronic pain(34). The primary outcome was the Visual Analog Scale (VAS) score, assessed at baseline, post-treatment, and follow-up. We calculated both the change score in VAS and clinical response as the primary outcomes. Change score (%) = [(VAS_post/follow-up_ -VAS_pre_) / VAS_pre_]×100%. Clinical response was defined as a ≥ 30% reduction in VAS relative to baseline(8, 24). Confidence intervals for response rates were calculated using the Wilson score method (binom package, R version 4.4.1).

Secondary outcome measures included the following:

(1) the Leeds Assessment of Neuropathic Symptoms and Signs (LANSS) score. (2) pain interference with daily activities, as measured by the Brief Pain Inventory (BPI). (3) the sensory and affective dimensions of pain, assessed by the Short-Form McGill Pain Questionnaire (SF-MPQ). (4) the Patient Global Impression of Change (PGIC), which was assessed at the post-treatment and follow-up timepoints. (5) changes in depressive and anxiety symptoms, evaluated by the Hamilton Depression Rating Scale (HAMD) and the Hamilton Anxiety Rating Scale (HAMA). (6) The patient’s overall health status, measured by the EQ-5D-5L questionnaire, and (7) brain functional connectivity—detailed in Sections 2.4.

### 2.7 Statistical analysis

All statistical analyses were conducted using R (version 4.4.1). Continuous data are presented as mean ± standard deviation (M ± SD). All statistical tests were two-tailed, and a significance level of α = 0.05 was adopted.

#### 2.7.1 Missing data handling

Multiple imputation (MI) was employed, to prevent sample size reduction and potential statistical bias arising from missing data. This procedure was implemented using the mice package in R. We generated 10 complete datasets using the predictive mean matching (pmm) algorithm (35, 36). The imputation model incorporated baseline demographics (e.g., age, sex) and all longitudinal continuous scale data. Categorical outcomes, including responder status and PGIC, were also imputed using the pmm algorithm. This approach allowed us to analyze the data based on the Intention-to-Treat (ITT) principle (N=20). All visualizations involving follow-up data (e.g., Sankey diagrams) and reported response rates were derived from this imputed ITT dataset. All subsequent statistical analyses were performed independently on each of the 10 imputed datasets, and the results were then pooled. Furthermore, to ensure that our conclusions were not dependent on imputation assumptions, a sensitivity analysis utilizing only the observed cases (Complete Case Analysis, n=18) was pre-specified for the primary outcome.

#### 2.7.2 Repeated measures ANOVA

To evaluate the longitudinal changes in outcome measures (i.e., VAS, SF-MPQ, BPI, LANSS, EQ-5D-5L, HAMD, and HAMA) across three time points—we conducted a repeated measures analysis of variance (ANOVA).

This analysis was performed separately for each of the 10 imputed datasets. For each dataset, a linear model was constructed with the scale score as the dependent variable, and time and subject ID as independent variables. The ANOVA results from the 10 models were then pooled using Rubin’s Rules, as implemented by the ‘D1’ function in the mice package, to yield a single, overall F-statistic and corresponding p-value for the main effect of time.

If a significant main effect of time (p < 0.05) was observed, pairwise comparisons were then performed using the ‘emmeans’ package to examine differences between time points. These comparisons were also conducted on each imputed dataset, and the results were subsequently pooled. To control for the family-wise error rate, p-values for the pairwise comparisons were adjusted using the Bonferroni method.

#### 2.7.3 Correlation analyses

To explore the neural correlates of clinical improvement, we examined the associations between treatment-induced changes in functional connectivity (ΔFC) and improvements across distinct clinical domains. Change scores (Δ) for functional connectivity and all clinical measures were computed as post-treatment minus pre-treatment values (Post – Pre).

Specifically, Pearson correlation analyses were conducted between ΔFC and changes in: (1) pain intensity (VAS), (2) neuropathic pain qualities (SF-MPQ), (3) pain interference (BPI), (4) health-related quality of life (EQ-5D-5L), (5) depressive symptoms (HAMD), and (6) anxiety symptoms (HAMA). All analyses were performed in MATLAB R2022b. For neuropathic pain signs, both the SF-MPQ and LANSS were administered. However, given that these two instruments assess the same underlying construct, only the SF-MPQ was entered into the primary correlation model to avoid redundancy. Thus, a total of six clinical domains were tested. Given the multiple comparisons (six tests), we applied false discovery rate (FDR) correction to control for Type I errors.

## RESULTS

### 3.1 Demographic and clinical characteristics

Thirty-two individuals were screened, resulting in a final cohort of 20 patients (exclusion reasons: VAS < 30, n=3; declined participation, n=9). Although all 20 patients completed the treatment protocol and post-treatment assessments, two were lost at the 1-month follow-up due to loss of contact (Figure 2) (Table 1). The cohort had a mean age of 60.0 ± 12.86 years and had been experiencing the current pain episode for an average of 2.83 ± 2.59 years. The sample was balanced in gender (9 males, 45.0%). The predominant diagnosis was postherpetic neuralgia (PHN; 60.0%), with the thoracic region being the most commonly affected area (45.0%).

**Figure 2.**
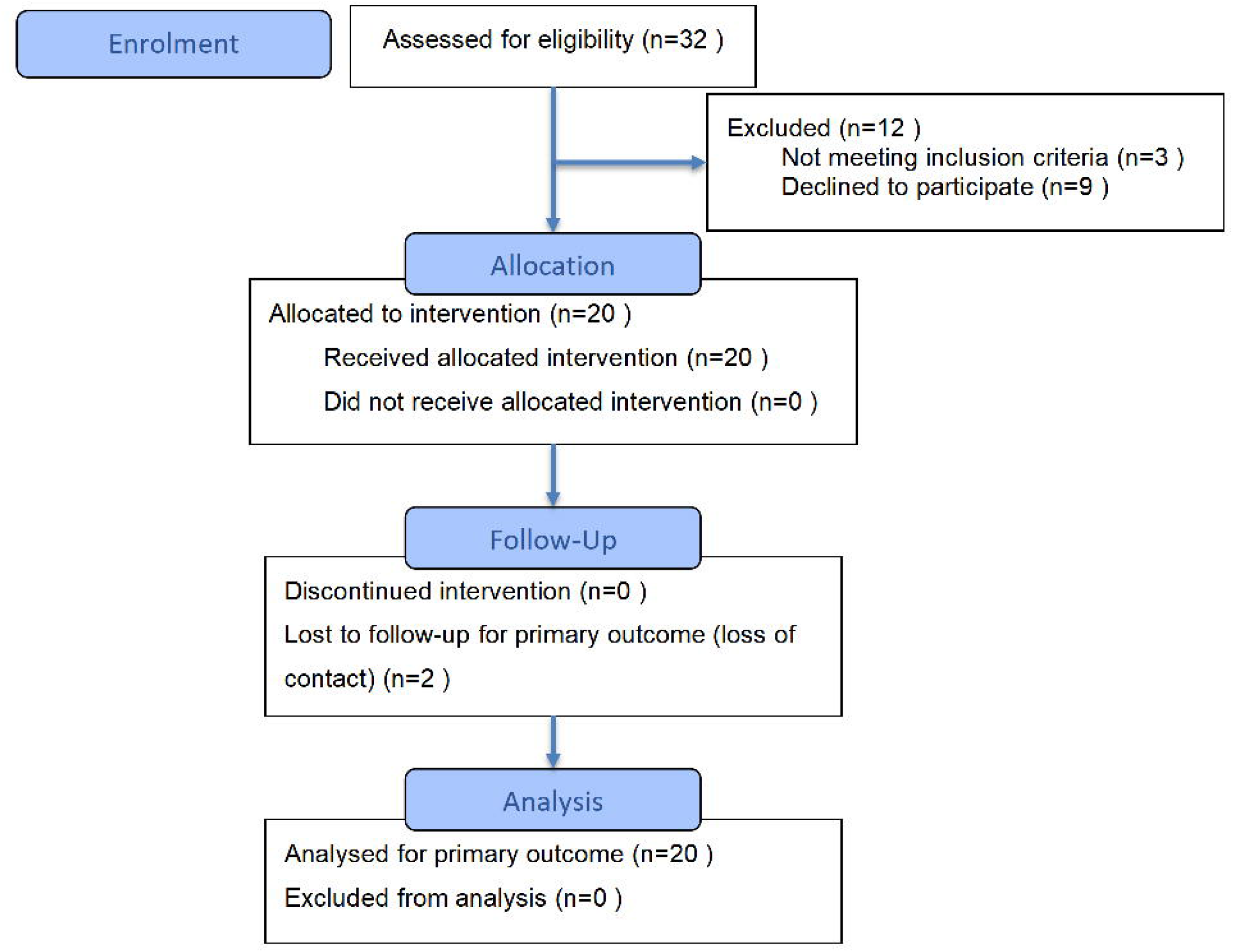
Flow diagram of the open-label trial. Multiple imputation was used to address missing data from 2 participants lost to follow-up.

Regarding the spatial distribution of the individualized SCAN targets, the group-average centroid (after mirroring to the left hemisphere) was located at MNI coordinates [-37.32, -13.22, 49.78]. The mean Euclidean distance between the individualized targets and this group centroid was 15.36 ± 13.27 mm, with individual distances ranging from 3.73 mm to 45.95 mm, indicating substantial spatial dispersion.

### 3.2 Treatment effects on the primary outcome

A significant time effect in pain severity was observed (F_(2, 34.15)_ = 6.77, p = .003). VAS scores decreased from baseline (50.2 ± 11.9) to post-treatment (36.7 ± 19.04, p = .001, Cohen’s d = 1.12, 95% CI [0.30, 1.94]) and follow-up (41.25 ± 20.24, p =.027, Cohen’s d = 0.85, 95% CI [0.02, 1.67]). The difference between post-treatment and follow-up was not significant (p = .680) (Figure 3a).

**Figure 3.**
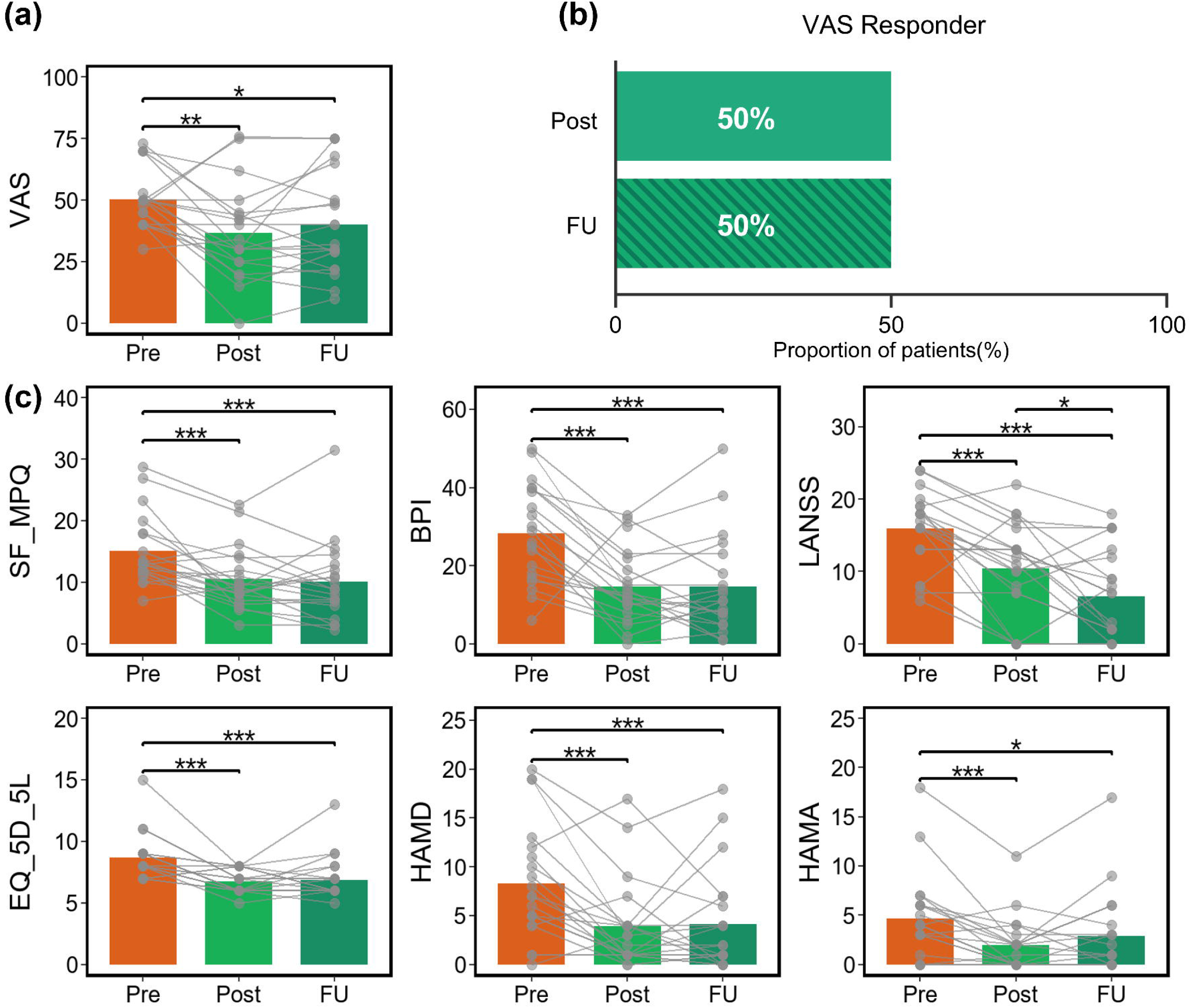
Clinical Outcomes. (a) Trajectories of the primary outcome, pain intensity, measured by the Visual Analog Scale (VAS) across time points. (b) The percentage of patients achieving clinical response at post-treatment and follow-up. Clinical response was defined as a ≥ 30% reduction in VAS scores from baseline. (c) Changes in secondary clinical measures, including pain interference, disability, and mood symptom scores. * indicates p < .05, ** indicates p < .01 and *** indicates *p* < .001

The response rate was 50.0% at the post-treatment (10/20; 95% Wilson CI: 29.92% – 70.07%) and 50.0% at the follow-up timepoint (10/20; 95% Wilson CI: 29.92% – 70.07%) (Figure 3b and Figure 4a). However, the response profiles varied across disease types. Patients with diabetic peripheral neuropathy (DPN) showed the highest response rate, with 100% (3/3) of patients classified as responders at both timepoints. Postherpetic neuralgia (PHN) showed a moderate response with a delayed therapeutic onset; the response rate increased from 41.7% (5/12) at post-treatment to 50.0% (6/12) at follow-up. Specifically, two non-responders converted to responders, loyed an open-label design to assess while one initial responder experienced symptom regression. Trigeminal neuralgia (TN) exhibited a stable response rate of 50% (1/2). In contrast, Traumatic neuropathy showed a lack of sustained benefit, where the initial response rate of 33.3% (1/3) dropped to 0% at follow-up.

**Figure 4.**
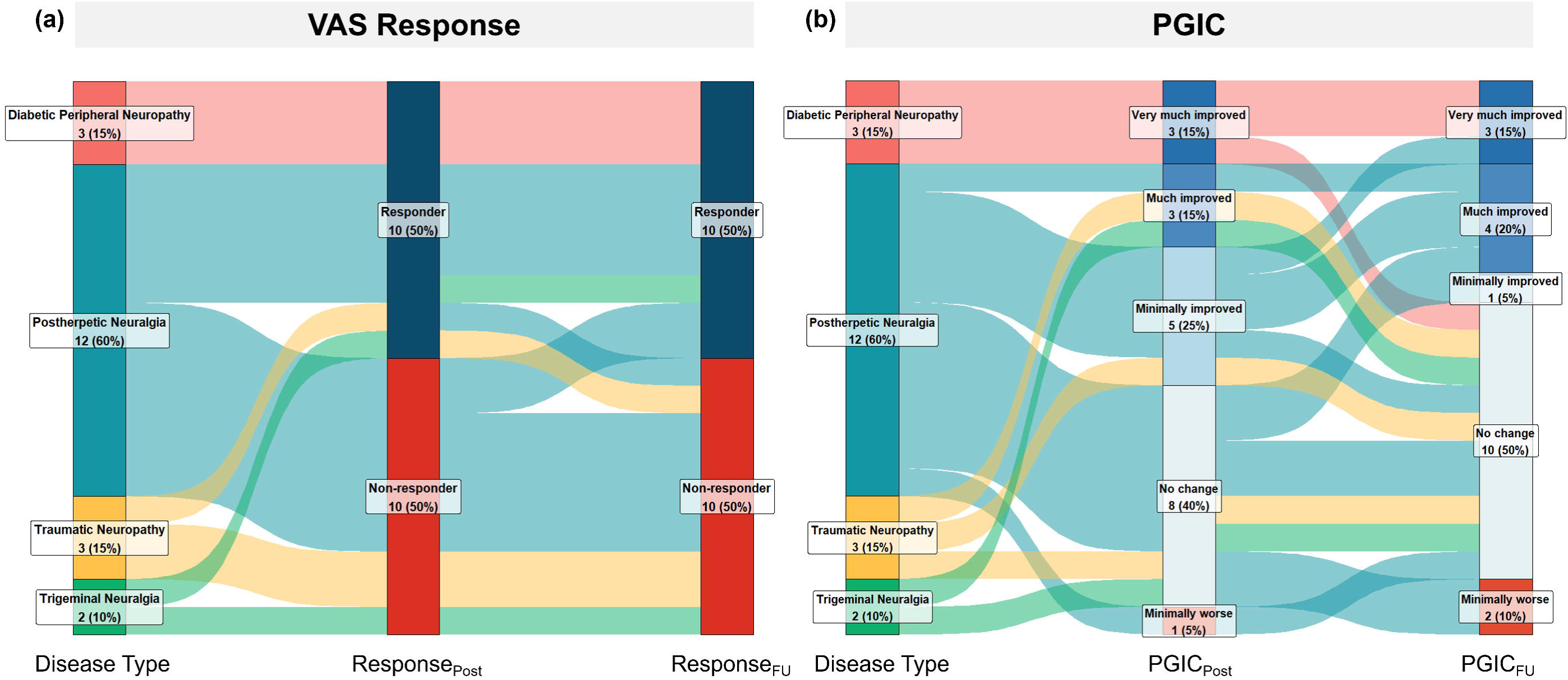
Change in Clinical Response (a) and PGIC (b) across different neuropathic pain types from post-treatment to follow-up. These Sankey diagrams visualize the flow and distribution of patient status across different time points. From left to right, the three columns of vertical nodes represent: (1) baseline disease type; (2) post-treatment status; and (3) follow-up status. The width of the flows is proportional to the number of patients in each path. Colour coding corresponds to the different disease types. Abbreviation: PGIC, Patients’ Global Impression of Change.

Importantly, the sensitivity analysis utilizing only the 18 observed cases yielded highly consistent results (main effect of time: F_(1.40, 23.71)_ =5.99, p = 0.014) with a clinical response rate of 50.0% (9/18) at post-treatment and 50.0% (9/18) at the 1-month follow-up. This confirms that the sustained therapeutic effect was robust and not driven by the multiple imputation model.

### 3.3 Treatment effects on the secondary outcomes

#### Short-Form McGill Pain Questionnaire(SF-MPQ)

SF-MPQ showed a significant main effect of time (F_(2,34.15)_ = 11.75, p < .001). SF-MPQ scores decreased from pre-treatment (15.07 ± 5.77) to post-treatment (10.52 ± 5.06, p= .001, Cohen’s d = 1.29, 95% CI [0.45, 2.13]) and follow-up (9.98 ± 6.57, p< .001, Cohen’s d = 1.40, 95% CI [0.52, 2.28]).

#### Brief Pain Inventory (BPI)

There was a significant main effect of time on BPI (F_(2,34.15)_ = 16.27, p < .001). BPI scores decreased from pre-treatment (28.25 ± 12.57) to post-treatment (14.65 ± 9.4, p < .001, Cohen’s d = 1.59, 95% CI [0.71, 2.47]) and follow-up (14.75 ± 12.77, p < .001, Cohen’s d = 1.58, 95% CI [0.67, 2.48]).

#### Leeds Assessment of Neuropathic Symptoms and Signs (LANSS)

Neuropathic symptom scores showed a significant main effect of time (F_(2,34.15)_ = 19.73, p < .001). Notably, the LANSS score exhibited a continuous and significant decline from pre-treatment (15.9 ± 5.32) to post-treatment (10.40 ± 6.56, p < .001, Cohen’s d = 1.21, 95% CI [0.38, 2.04]), and further significantly decreased from post-treatment to follow-up (6.75 ± 6.46, p = .030, Cohen’s d = 0.84, 95% CI [0.01, 1.66]).

#### EQ-5D-5L Quality of Life

Quality of life, indicated by lower scores, significantly improved (F_(2,34.15)_ = 17.45, *p* < .001). Scores decreased from pre-treatment (8.70±1.87) to post-treatment (6.75 ± 0.97, *p* < .001, Cohen’s d = 1.67, 95% CI [0.78, 2.56]) and follow-up (6.90 ± 1.71, p < .001, Cohen’s d = 1.60, 95% CI [0.69, 2.51]). The improvement was sustained, with no significant difference between post-treatment and follow-up (p = .97).

#### Hamilton Depression Rating Scale (HAMD-17)

Depression scores significantly reduced across time (F_(2,34.15)_ = 9.93, *p* < .001). Scores at post-treatment (3.90± 4.61, p < .001, Cohen’s d = 1.27, 95% CI [0.43, 2.11]) and follow-up (4.50 ± 5.12, p = .001, Cohen’s d = 1.19, 95% CI [0.34, 2.05]) were significantly lower than in pre-treatment (8.30 ± 5.92). There was no significant difference between post-treatment and follow-up (p = .97).

#### Hamilton Anxiety Rating Scale (HAMA-14)

Anxiety scores also showed a significant main effect of time (F_(2,34.15)_ = 9.13, p < .001). Scores significantly decreased from pre-treatment (4.65 ± 4.45) to post-treatment (1.95 ± 2.74, p < .001, Cohen’s d = 1.33, 95% CI [0.48, 2.17]) and follow-up (2.65 ± 4.25, p = .020, Cohen’s d = 0.88, 95% CI [0.06, 1.71]). No significant difference was found between post-treatment and follow-up (p= .36).

#### Patient Global Impression of Change (PGIC)

The Patient Global Impression of Change (PGIC) was assessed at post-treatment and follow-up. At post-treatment, 55% of patients reported global improvement, stratified as “Very Much Improved” (15%), “Much Improved” (15%), and “Minimally Improved” (25%). However, the improvement declined to 40% at the 1-month follow-up (“Very Much Improved”: 15%; “Much Improved”: 20%; “Minimally Improved”: 5%). Notably, all three patients with diabetic peripheral neuropathy (DPN) reported a significant improvement in their pain symptoms at the post-treatment assessment.

Regarding no-response or deterioration, the proportion of patients reporting “No Change” rose from 40% post-treatment to 50% at follow-up. Similarly, the incidence of symptom worsening increased from 5% to 10% over the same period(Figure 4b).

### 3.4 Changes in functional connectivity and associations with clinical improvements

We calculated the functional connectivity between the SCAN and CON networks. There was no significant difference in functional connectivity when comparing the pre-treatment with post-treatment (0.43 ± 0.23 vs.0.40 ± 0.20, p = .547). Correlational analysis revealed that changes in functional connectivity were significantly negatively associated with improvements in HAMA (r = -.565, uncorrected p = .009, FDR-corrected p = .036), HAMD (r = -.552, uncorrected p = .012, FDR-corrected p = .036), and BPI scores (r = -.503, uncorrected p = .024, FDR-corrected p = .048) (Figure 5).

**Figure 5.**
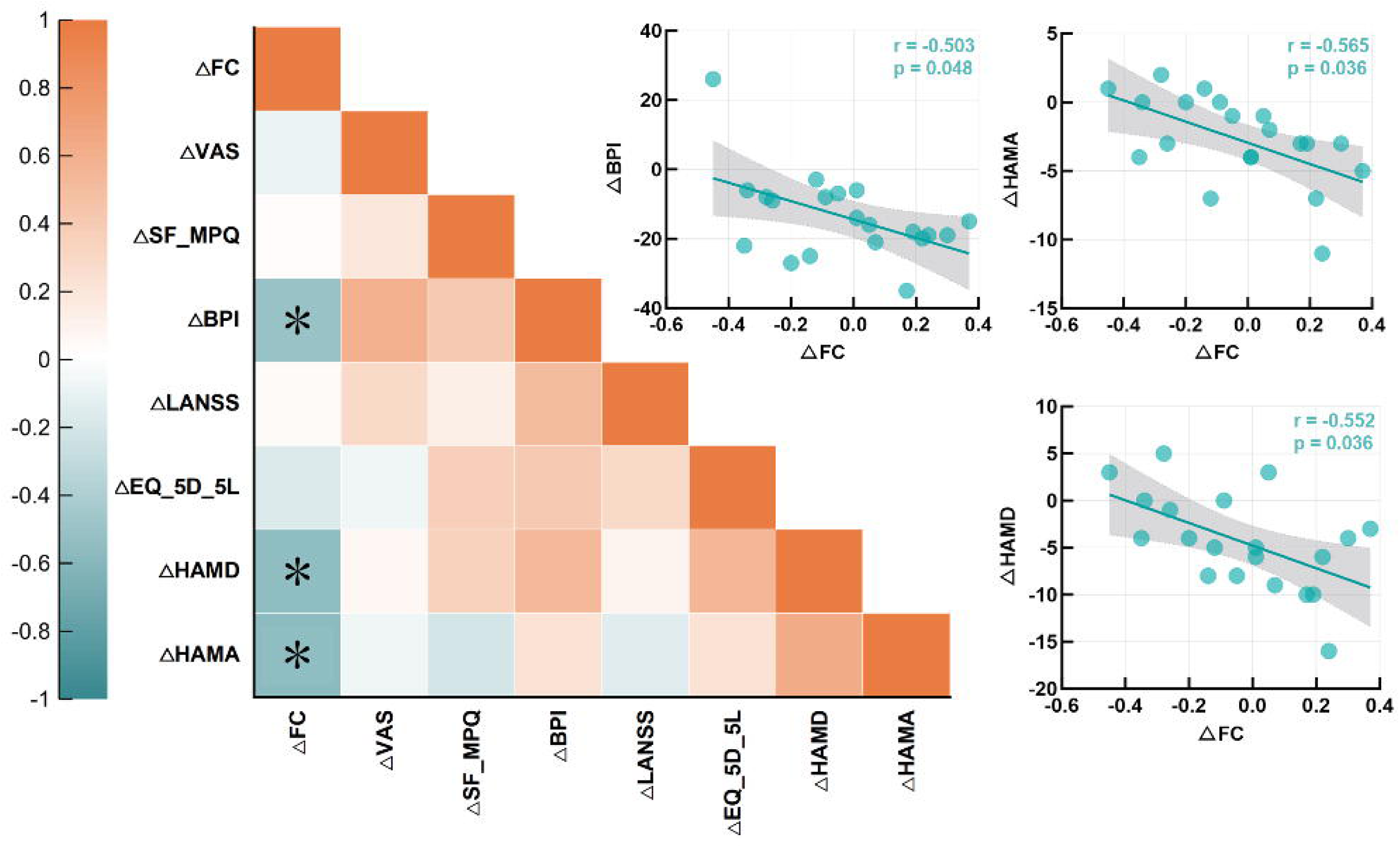
Relationships between SCAN-CON functional connectivity and clinical outcomes. Heatmap displaying Pearson correlation coefficients (r) between SCAN-CON functional connectivity values and clinical measures. Orange and green cells denote positive and negative correlations, respectively. Notably, SCAN-CON connectivity changes were directly associated with pain interference, depressive, and anxiety symptoms, but not with pain intensity. *Significance levels: \**p* < .05 (FDR corrected).

A sensitivity analysis on observed cases only (n=18) confirmed the robustness of these findings. The ΔFC-HAMA association remained significant after FDR correction (r =−.655, uncorrected p =.003, FDR-corrected p =.018). Although associations for HAMD (r =−.542, uncorrected p =.020, FDR-corrected p =.060) and BPI (r =−.490, uncorrected p =.039, FDR-corrected p =.078) did not survive FDR correction in this smaller sample, they maintained consistent effect sizes and statistical trends, indicating the primary results were stable and not artifacts of data imputation.

### 3.5 Safety and tolerability

No serious side effects of rTMS were noted. Minor adverse effects were noted in five patients, including fatigue (10%) and dizziness (15%) (see Table 1).

## DISCUSSION

Leveraging the newly defined CON-SCAN network, this open-label clinical trial was the first connectivity-guided personalised rTMS trial for neuropathic pain. Our rTMS treatment achieved a response rate of 50%. Analgesic efficacy was maintained until 1-month follow-up. Moreover, exploratory descriptive observations suggested variable response profiles across different neuropathic pain etiologies, though these require validation in larger cohorts. In addition, increased CON-SCAN connectivity following TMS therapy was associated with a larger improvement in certain pain experiences.

A commonly adopted rTMS protocol for neuropathic pain is 10 sessions over 2 weeks (10-Hz, ∼1,500 pulses in each session) (6, 7, 9, 10). Using this paradigm, previous studies reported a ∼40% response rate at the end of treatment in postherpetic neuralgia (9, 10). In line with these 2 trials, 60% of our participants were individuals with postherpetic neuralgia. With personalised targeting, we observed a response rate of 50% at the end of treatment. More importantly, this efficacy was maintained at 1-month follow-up. Despite these promising results, the clinical efficacy observed here reflects considerable inter-individual variability, with the mean VAS reduction falling slightly below our predefined 30% threshold for individual clinical response. While the reduction in VAS scores was statistically significant, the average decrease of approximately 27% indicates a moderate overall treatment effect. In addition, the 50% responder rate in our pilot sample (N = 20) was associated with broad 95% confidence intervals (29.9%–70.1%), highlighting substantial uncertainty. Thus, our findings should be interpreted modestly as encouraging yet variable signals of therapeutic efficacy, rather than evidence of a uniformly robust clinical effect. These benefits may be associated with network-optimised targeting, in contrast to previous studies where this was mainly determined by motor response output or electroencephalography (EEG) 10-20 system. Given the open-label design, these findings provide preliminary evidence supporting the feasibility and tolerability of personalised rTMS targeting for neuropathic pain, highlighting promising clinical signals that require future validation in randomized, sham-controlled trials.

Our study also provides novel data on the treatment effects on subtypes of neuropathic pain. A multicentre, large trial revealed that M1-rTMS was effective for neuropathic pain caused by traumatic/surgical nerve lesion or sensory polyneuropathy (37). Other trials have also confirmed a positive effect of M1-rTMS on trigeminal neuralgia (38) and postherpetic neuralgia (PHN) (6, 9, 10). These subtypes of neuropathic pain are represented by the majority of our patients (15/20), and our findings support the benefits of M1-rTMS treatment for these conditions. We also noted descriptive variations in treatment response across different etiologies, such as all three DPN patients achieving clinical response. However, we strongly emphasize that with sub-cohorts as small as N=3, these subtype-specific findings represent exploratory descriptive observations that are highly hypothesis-generating. Whether this pattern reflects a true differential treatment effect or merely chance variation within a small subgroup cannot be determined from the present dataset. Investigation in a larger, etiology-stratified cohort would be necessary to assess whether DPN responds differently to this personalised approach.

The clinical presentation of neuropathic pain extends beyond the sensory dimension to include significant affective/emotional, and cognitive components, which collectively contribute to the overall burden of the disease (39, 40). Beyond pain intensity, our personalised rTMS treatment also demonstrated consistent effects on other pain experiences, such as pain symptoms, pain interference, negative emotions, and overall health status. These benefits were also maintained to 1 month after the treatment. Moreover, neuropathic symptoms and sensory signs assessed by the LANSS examination demonstrated a further improvement from post-treatment to follow-up. Together, these positive findings provide support for the effectiveness of personalised rTMS treatment for neuropathic pain. They also highlight the multidimensional nature of pain experiences (41, 42). However, findings regarding mood and anxiety improvements must be interpreted with strict caution. By design, patients with severe psychiatric disorders were excluded from this trial, resulting in a low baseline symptom burden for both depression (mean HAMD ∼8.30) and anxiety (mean HAMA ∼4.65). Therefore, these results should not be extrapolated to imply broad antidepressant or anxiolytic efficacies of the SCAN-guided protocol in patients with primary mood disorders.

In addition to clinical efficacy, we further provided evidence that CON-SCAN network was associated with rTMS treatment effect in neuropathic pain. Increased CON-SCAN connectivity following the course of rTMS was associated with a larger improvement in pain interference, depression, and anxiety, but notably not with reductions in pure pain intensity. These findings indicate that clinical efficacy was associated with parallel changes in neural connectivity metrics. This dissociation aligns perfectly with our theoretical framework. The CON-SCAN network is widely recognized for encoding the affective-attentional and cognitive dimensions of pain, rather than ascending nociception alone. Therefore, these results suggest that the therapeutic value of this personalized targeting strategy extends beyond pure analgesia. Consequently, the primary clinical contribution of this CON-SCAN-guided approach should be framed as facilitating robust ’multidimensional symptom relief. This likely reflects the role of CON in modulating the affective dimensions of pain. Despite these associations, it is interesting to note that statistically significant changes in CON-SCAN connectivity were not identified in our data. We acknowledge that this absence of a universal network shift limits our ability to make strong mechanistic claims. While this lack of a group effect might relate to sample heterogeneity or individual-specific rebalancing of connectivity, such explanations remain speculative without larger cohorts. Therefore, our results should be interpreted with caution: our data demonstrate that individual variations in network connectivity scale with symptom relief, but they do not support a generalized, group-level reorganization of the SCAN-CON network. Overall, this is an exploratory mechanistic support but not validation.

Beyond the CON-SCAN network, other neurocircuitries may have also played a role in mediating the analgesic effects of M1-rTMS. A potential mechanism involves the engagement of the descending pain modulatory system, which hinges on key structures like the periaqueductal grey (PAG) and thalamus (43). This is evidenced by the association between thalamocortical tract integrity and the analgesic efficacy of M1-rTMS (44). Specifically, neuropathic pain caused by lesions over the brainstem and thalamic regions benefited less from M1-rTMS, suggesting that the descending pain modulatory system is critical for M1-rTMS to generate analgesia (38). In addition, the presence of long-term effects in our data and others (37) also indicate neuroplastic changes induced by repetitive sessions of rTMS. Indeed, a series of studies have reported neuroplastic changes following motor cortex rTMS in neuropathic pain (45–48).

The optimal person-specific stimulation target was computed within the spatial bounds of the SCAN mask. We prioritized the SCAN as the primary stimulation site due to its cortical accessibility for standard figure-of-eight coils, which allow for high focal precision. While the CON contains key nodes involved in pain processing (e.g., dACC and insula)(19), these regions are located in deeper cortical structures. Stimulating these areas would require specialized deep-TMS coils (e.g., H-coils or double-cone coils), which often result in significantly reduced focal specificity and may recruit unintended neighboring regions(49). Therefore, targeting the SCAN—a surface-accessible network with strong functional coupling to the CON—provides a more controlled and precise neuromodulatory approach. Furthermore, our spatial analysis robustly justifies the necessity of personalized targeting over a generic group-level approach. As reported, the individualized targets exhibited vast spatial dispersion from the group centroid (mean deviation >15 mm). Because the peak electric field of a standard figure-of-eight coil drops off sharply within 5–10 mm, this magnitude of spatial variance confirms that individualized targets did not simply cluster in a narrow common region. Relying on a simpler group-average target would have structurally missed the optimal functional connectivity peak in the majority of patients, thereby highlighting the indispensable spatial added value of our network-guided approach. However, established precision functional mapping studies have confirmed that sensorimotor network topography exhibits exceptionally high within-subject stability over time(50), providing strong theoretical support for the robustness of our individualized targeting strategy.

We emphasize that this open-label pilot study lacked a conventional M1-targeting comparator arm and was therefore not designed to test whether personalised targeting enhances efficacy relative to standard approaches. The observed clinical improvements cannot be disentangled from nonspecific treatment effects. A randomised controlled trial incorporating both sham and active conventional-target arms is required to definitively evaluate the added value of SCAN-guided personalisation. Such a trial represents the logical next step of this research program and is currently being planned.

There were some limitations in this study. Firstly, we employed an open-label design to assess the efficacy of this approach without a traditional control group. While we recognize that the absence of a control group limits the generalizability of the findings and makes it difficult to exclude placebo effects, the results still provide valuable preliminary evidence for the efficacy of personalized CON-SCAN network-targeted rTMS in neuropathic pain. Future studies should incorporate randomized controlled trials (RCTs) to further validate the effectiveness and mechanisms of this approach. We anticipate that the efficacy of this targeted TMS therapy can now be further improved using higher dose and more accelerated TMS therapy. Protocols such as Stanford Neuromodulation Therapy (51), and our own variations on this (52, 53), have demonstrated substantial advances in clinical efficacy. Next, it is conceivable that precision functional mapping (PFM) of individual network architecture could further improve TMS therapeutic outcomes. However, PFM conventionally requires at least 30-60 minutes of resting state data (50). Moreover, the majority of PFM research is based on several hours of scan data (19). Acquiring several hours of scan data is unrealistic for patients suffering from pain. Importantly, SCAN (and CON) have been delineated in large cohorts based on short scans and these network map templates are more readily implemented (19). Relatedly, further research should investigate the contribution of personalised vs group-average targeting strategies here. We note that many participants’ targets were ultimately similar in their position around the central inter-effector region, and that this region is fairly constrained. There are other neurocircuitries that can be explored to optimise treatment efficacy for neuropathic pain (16, 17). Nonetheless, the present work focuses on optimisation within one of the major TMS targets for pain, namely the motor cortex.

Our sample included several common subtypes of neuropathic pain. Due to a small sample size in each subtype, it is difficult to draw strong conclusions of different effects on these subtypes. Our data identified a stronger effect in DPN than other subtypes, which opens discussions for future studies to validate these effects with large samples. Although we used connectivity and neuronavigation to pinpoint an optimal target, it was not adjusted by e-field modelling. A recent trial has adopted this procedure to improve rTMS induced electric field (54). Due to ethical consideration, patients maintained their medication regimen during the treatment, as is common for rTMS studies in neuropathic pain (37). This might interact with rTMS treatment, and future studies with a sham treatment could examine this effect. Regarding the observed neuroimaging-clinical correlations—which exhibited medium effect sizes (r ≥ 0.5) and survived false discovery rate (FDR) correction for multiple comparisons—these associations provide preliminary evidence linking CON-SCAN connectivity changes to clinical improvement. While the exact causal relationship between these brain changes and clinical improvements remains to be fully understood, the findings suggest that functional changes in brain networks may be associated with improvements in pain, anxiety, and depression. Further research is needed to validate these mechanisms in larger, controlled trials.

To conclude, this is the first study to investigate personalised connectivity based rTMS treatment for neuropathic pain. By moving beyond a “one-size-fits-all” approach, this personalised therapeutic approach achieved a 50% response rate and maintained this analgesic efficacy to 1-month follow-up. Crucially, the significant association identified between neurophysiological connectivity shifts and clinical symptom relief is consistent with the CON-SCAN network as a candidate therapeutic target, providing preliminary support for the mechanistic rationale underlying this personalized targeting approach. These findings represent a significant step toward precision neuromodulation in chronic pain management and provide a compelling foundation for future large-scale, double-blind randomised controlled trials to establish definitive clinical efficacy.

## Ethics approval and consent to participate

All participants in this study enrolled voluntarily and signed an informed consent form prior to the official initiation of the study. This study has obtained approval from the Ethics Committees of the Affiliated Hospital of Hangzhou Normal University (Ethics No.: 2024-E2-HS-008) and was conducted in strict compliance with the requirements of the Code of Ethics of the World Medical Association (Declaration of Helsinki). The trial was registered with the Chinese Clinical Trial Registry: ChiCTR2400094568 (Registered 2024-12-24, “Functional MRI connectivity guided personalised targeting to optimise rTMS treatment efficacy in neuropathic pain conditions”).

## Consent for publication

Written informed consent for publication of their clinical details was obtained from the patient. A copy of the consent form is available for review by the Editor of this journal.

## Data Availability

All the data would be available upon reasonable request from the corresponding author.

## Authors’ contributions

ZH, AZ, RC, XC and ZF contributed to study design, data analysis, and writing-up. YL, SW and HL participated in data collection. All authors read and approved the final manuscript.

## Acknowledgements

Not applicable

## List of abbreviations

Abbreviation Full Meaning

AC: Anterior Commissure
ACC: Anterior Cingulate Cortex
ANOVA: Analysis of Variance
BOLD: Blood-Oxygen-Level-Dependent
BPI: Brief Pain Inventory
CON: Cingulo-Opercular Network
DLPFC: Dorsolateral Prefrontal Cortex
DPN: Diabetic Peripheral Neuropathy
EPI: Echo-Planar Imaging
EQ-5D-5L: 5-level EuroQol 5-dimension questionnaire
FDR: False Discovery Rate
fMRI: Functional Magnetic Resonance Imaging
FOV: Field of View
FWHM: Full Width at Half-Maximum
HAMA: Hamilton Anxiety Rating Scale
HAMD: Hamilton Depression Rating Scale
HCP: Human Connectome Project
IASP: International Association for the Study of Pain
IMMPACT: Initiative on Methods, Measurement, and Pain Assessment in Clinical Trials
LANSS: Leeds Assessment of Neuropathic Symptoms and Signs
M1: Primary Motor Cortex
MEP: Motor-Evoked Potential
MI: Multiple Imputation
MMSE: Mini-Mental State Examination
MRI: Magnetic Resonance Imaging
NIBS: Non-invasive Brain Stimulation
NP: Neuropathic Pain
PAG: Periaqueductal Grey
PFM: Precision Functional Mapping
PGIC: Patient Global Impression of Change
PHN: Postherpetic Neuralgia
RMT: Resting Motor Threshold
ROI: Region of Interest
rTMS: Repetitive Transcranial Magnetic Stimulation
SCAN: Somato-Cognitive Action Network
SF-MPQ: Short-Form McGill Pain Questionnaire
SGC: Subgenual Cingulate Cortex
SNR: Signal-to-Noise Ratio
TE: Echo Time
TI: Inversion Time
TMS: Transcranial Magnetic Stimulation
TN: Trigeminal Neuralgia
TR: Repetition Time
VAS: Visual Analogue Scale

