## Supplementary material for "Personalising Transcranial Magnetic Stimulation Therapy for Neuropathic Pain with Somato-Cognitive Action Network Connectivity to Cingulo-Opercular Network: A Preliminary Open-Label Study": Table 1

| Table 1. Demographic and clinical information (N=20) | |
| --- | --- |
|  | Mean ± SD / Number(%) |
| **Demographic variables** |  |
| Age (years) | 60.00 ± 12.86 |
| Height (cm) | 163.05 ± 8.86 |
| Weight (kg) | 59.95 ± 9.76 |
| Duration of current pain episode (years) | 2.83 ± 2.59 |
| Gender(male)^a^ | 9 (45.0%) |
| Education |  |
| Primary or below | 6 (30.0%) |
| Lower secondary | 10 (50.0%) |
| Upper secondary | 2 (10.0%) |
| Tertiary education | 2 (10.0%) |
| **Classification^a^** |  |
| Postherpetic neuralgia (PHN) | 12 (60.0%) |
| Painful diabetic peripheral neuropathy (DPN) | 3 (15.0%) |
| Post-traumatic neuralgia | 3 (15.0%) |
| Trigeminal neuralgia (TN) | 2 (10.0%) |
| **TMS variables** |  |
| Resting motor threshold (% of maximum stimulator output) | 60.55 ± 11.96 |
| **Drug intake^a^** |  |
| Antiepileptic & Neuropathic Pain Agents |  |
| Pregabalin | 18 (90.0%) |
| Gabapentine | 2 (10.0%) |
| Miloxagabine | 1 (5.0%) |
| Carbamazepine/Oxcarbazepine | 3 (15.0%) |
| Analgesics |  |
| Weak Opioids (e.g., Tramadol, Oxycodone/Acetaminophen) | 2 (10.0%) |
| Psychotropic Agents |  |
| Duloxetine | 2 (10.0%) |
| Other (Agomelatine) | 1 (5.0%) |
| Adjunctive Therapies |  |
| Local Anesthetics (e.g., Lidocaine) | 2 (10.0%) |
| Vitamin B12/ Huoxue Zhitong Jiaonang | 2 (10.0%) |
| **Hypertension Status^a^** |  |
| Well-controlled hypertension | 5 (25.0%) |
| No hypertension | 15 (75.0%) |
| **Nerves involved in the case^a^** |  |
| Head and Face (incl. Trigeminal) | 4 (20.0%) |
| Neck (Cervical) | 1 (5.0%) |
| Chest (Thoracic) | 9 (45.0%) |
| Lumbar Region | 1 (5.0%) |
| Lower Limbs (Legs/Feet) | 2 (10.0%) |
| **Adverse Effects^a^** |  |
| Fatigue | 2(10.00%) |
| Dizziness | 3(15.00%) |
| No | 15(75.00%) |

Note. Items marked with superscript *a* denote categorical variables.
