## Supplemental Table 1 for "Personalising Transcranial Magnetic Stimulation Therapy for Neuropathic Pain with Somato-Cognitive Action Network Connectivity to Cingulo-Opercular Network: A Preliminary Open-Label Study"

| Supplement Table.1 Participant Information | | | | | | | | | | | | | | | | |
| --- | --- | --- | --- | --- | --- | --- | --- | --- | --- | --- | --- | --- | --- | --- | --- | --- |
| **ID** | **Gender** | **Classification** | **Nerves involved in the case** | **Stimulation side** | **RMT** | **Native** | | | **Standard** | | | **VAS** | | | **Post-treatment responder** | **Follow‑up VAS responder** |
|  |  |  |  |  |  | **X** | **Y** | **Z** | **X** | **Y** | **Z** | **pre** | **post** | **FU** |  |  |
| sub01 | male | PHN | Thoracic | R | 70 | 52.89 | -8.49 | 20.09 | 61.19 | 0.09 | 23.19 | 50 | 42 | 68 | 0 | 0 |
| sub02 | female | PHN | Thoracic | L | 46 | -39.52 | -16.12 | 42.22 | -44.44 | -7.53 | 48.49 | 40 | 30 | 30 | 0 | 0 |
| sub03 | male | Post-traumatic neuralgia | Lumbar/Groin | R | 40 | 62.71 | 6.33 | 9.44 | 60.19 | 5.5 | 14.6 | 40 | 25 | 30 | 1 | 0 |
| sub04 | male | PHN | Head and Face | L | 55 | -10.2 | -52.98 | 45.95 | -13.07 | -29.56 | 63.01 | 40 | 40 | 40 | 0 | 0 |
| sub05 | female | PHN | Thoracic | L | 47 | -41.75 | -26.09 | 40.44 | -42.01 | -10.94 | 55 | 40 | 30 | 40 | 0 | 0 |
| sub06 | female | TN | Head and Face | L | 61 | -35.64 | -20.03 | 39.52 | -37.34 | -9.82 | 51.79 | 50 | 50 | 65 | 0 | 0 |
| sub07 | female | PHN | Thoracic | L | 68 | -33.73 | -28.93 | 35.28 | -35.6 | -18.86 | 54.36 | 70 | 30 | 65 | 1 | 0 |
| sub08 | male | DPN | Lower Limbs (Feet) | L | 47 | -44.33 | -21.85 | 43.71 | -40.9 | -10.63 | 54.62 | 40 | 0 | 10 | 1 | 1 |
| sub09 | male | PHN | Head and Face | L | 68 | -33.67 | -33.32 | 29.94 | -36.2 | -17.49 | 39.96 | 70 | 45 | 48 | 1 | 1 |
| sub10 | male | DPN | Lower Limbs | L | 65 | -39.52 | -30.22 | 40.03 | -39.37 | -15.5 | 51.91 | 50 | 19 | 13 | 1 | 1 |
| sub11 | female | TN | Head and Face | R | 45 | 9.46 | -40.5 | 52.71 | 16.15 | -22.18 | 63.91 | 73 | 42 | 49 | 1 | 1 |
| sub12 | female | PHN | Thoracic | R | 70 | 29.21 | -19.36 | 36.71 | 36.88 | -14.99 | 40.71 | 45 | 20 | 22 | 1 | 1 |
| sub13 | female | Post-traumatic neuralgia | Lumbar | L | 68 | -42.6 | -20.7 | 39.44 | -42.29 | -10.68 | 50.65 | 48 | 76 | 75 | 0 | 0 |
| sub14 | female | PHN | Thoracic | L | 80 | -39.7 | -23.1 | 41.86 | -43.96 | -6.92 | 52.11 | 50 | 30 | 32 | 1 | 1 |
| sub15 | female | PHN | Thoracic | L | 60 | -19 | -36.4 | 48.98 | -17.74 | -25.6 | 61.58 | 45 | 15 | 29 | 1 | 1 |
| sub16 | male | PHN | Thoracic | L | 58 | -44.4 | -26.2 | 32.32 | -45.45 | -10.39 | 44.55 | 30 | 34 | 10 | 0 | 1 |
| sub17 | male | DPN | Lower Limbs (Legs) | R | 60 | 37.79 | -18.5 | 47.9 | 41.71 | -7.77 | 52.62 | 53 | 25 | 20 | 1 | 1 |
| sub18 | male | PHN | Cervical | L | 80 | -40.1 | -17.6 | 39.37 | -40.29 | -11.52 | 44.88 | 50 | 44 | 29 | 0 | 1 |
| sub19 | female | PHN | Head and Face | L | 74 | -18.8 | -33.37 | 70.63 | -17.96 | -29.25 | 74.53 | 50 | 75 | 75 | 0 | 0 |
| sub20 | female | Post-traumatic neuralgia | Head and Face | L | 49 | -30.2 | -31.7 | 34.14 | -33.74 | -10.44 | 53.23 | 70 | 62 | 50 | 0 | 0 |
